# Combination of Antidepressants and Antipsychotics as A Novel Treatment Option for Psychosis in Alzheimer’s Disease

**DOI:** 10.1101/2023.01.24.23284970

**Authors:** Peihao Fan, Lang Zeng, Ying Ding, Julia Kofler, Jonathan Silverstein, Joshua Krivinko, Robert A Sweet, Lirong Wang

**Affiliations:** Computational Chemical Genomics Screening Center, Department of Pharmaceutical Sciences/School of Pharmacy, University of Pittsburgh, Pittsburgh, USA; Graduate School of Public Health, University of Pittsburgh, Pittsburgh, USA; Division of Neuropathology, Department of Pathology, University of Pittsburgh Medical Center, Pittsburgh, USA; Department of Biomedical Informatics, School of Medicine, University of Pittsburgh, Pittsburgh, USA; Department of Psychiatry, University of Pittsburgh, School of Medicine, Pittsburgh, PA; UPMC Endowed Professor in Psychiatric Neuroscience, Professor of Neurology and Clinical and Translational Science, 3811 O’Hara Street, Pittsburgh, PA 15213; Computational Chemical Genomics Screening Center, Department of Pharmaceutical Sciences/School of Pharmacy, University of Pittsburgh, 6456 Salk Hall, 3501 Terrace Street, Pittsburgh, PA 15213

## Abstract

**Background:** Psychotic symptoms are reported as one of the most common complications of Alzheimer’s disease (AD), affecting approximately half of AD patients, in whom they are associated with more rapid deterioration and increased mortality. Empiric treatments, namely first and second-generation antipsychotics, confer modest efficacy in AD patients with psychosis (AD+P) and themselves increase mortality. A recent genome-wide meta-analysis and early clinical trials suggest the use and beneficial effects of antidepressants among AD+P patients. This motivates our rationale for exploring their potential as a novel combination therapy option amongst these patients.

**Methods:** We included University of Pittsburgh Medical Center (UPMC) electronic medical records (EMRs) of 10,260 AD patients from January 2004 and October 2019 in our study. Survival analysis was performed to assess the effects of the combination of antipsychotics and antidepressants on the mortality of these patients. To provide more valuable insights on the hidden mechanisms of the combinatorial therapy, a protein-protein interaction (PPI) network representing AD+P was built, and network analysis methods were used to quantify the efficacy of these drugs on AD+P. An indicator score combining the measurements on the separation between drugs and the proximity between the drugs and AD+P was used to measure the effect of an antipsychotic-antidepressant drug pair against AD+P.

**Results:** Our survival analyses replicated that antipsychotic usage is strongly associated with increased mortality in AD patients while the co-administration of antidepressants with antipsychotics showed a significant beneficial effect in reducing mortality. Our network analysis showed that the targets of antipsychotics and antidepressants are well-separated, and antipsychotics and antidepressants have similar proximity scores to AD+P. Eight drug pairs, including some popular recommendations like Aripiprazole/Sertraline and other pairs not reported previously like Iloperidone/Maprotiline showed higher than average indicator scores which suggest their potential in treating AD+P via strong synergetic effects as seen in our study.

**Conclusion:** Our proposed combinations of antipsychotics and antidepressants therapy showed a strong superiority over current antipsychotics treatment for AD+P. The observed beneficial effects can be further strengthened by optimizing drug-pair selection based on our systems pharmacology analysis.

## 1. Introduction

Approximately more than 50% of Alzheimer’s disease (AD) patients experience psychotic symptoms and other neuropsychiatric complications ^1^. AD patients with psychosis (AD+P) are considered a subgroup of patients who experience more severe symptoms including greater cognitive impairment and a quicker cognitive decline ^2^. AD+P is also associated with higher rates of co-occurring agitation ^3^, aggression ^3, 4^, depression ^5, 6^, mortality^7^, functional impairment ^8^, and increased caregiver burden ^9^ than AD patients without psychosis (AD-P).

Currently, no specific medication has been approved by FDA for AD+P. Second-generation antipsychotics (SGAs) have been widely used and recommended by geriatric experts in the management of psychosis in AD ^10-12^. However, their usage is greatly limited by increased risk of adverse events and the likelihood of co-morbid health problems ^13, 14^. This prompted the FDA to issue a “black-box” warning in 2005 to highlight the increased risk of mortality for patients with dementia who are treated with SGAs ^15^. Meanwhile, antipsychotics have demonstrated modest efficacy in treating psychosis, aggression, and agitation in individuals with dementia ^16-18^. This emphasizes the need to develop safer and more effective treatment options for AD+P.

Based on our previous studies, antipsychotics do not engage the underlying biology of AD+P, and therefore their modest effectiveness is unsuprising^19^. In order to identify safe and effective treatments for AD+P, it is essential to have a comprehensive understanding of the underlying biology of AD+P. A recent study reported that the heritability of psychosis in AD is estimated to be 61%, thus suggesting a strong association between AD+P and genetic variations^20^. Another study performed a large genome-wide meta-analysis on 12,317 AD subjects with or without psychosis ^21^. The authors reported that AD⍰+ ⍰P was not significantly genetically correlated with schizophrenia, but it was negatively correlated with bipolar disorder and positively correlated with depression. These associations provide a biologic rationale for repurposing antidepressant agents as novel treatment options for AD+P in our current study.

To answer if antidepressants are effective in managing neuropsychiatric symptoms in AD patients, nine clinical trials involving 692 patients were conducted. Five of them compared antidepressants with placebo and 4 compared with antipsychotics ^22-32^. However, only two selective serotonin reuptake inhibitors (SSRIs) sertraline (Zoloft) and citalopram (Celexa) were studied and the antipsychotics in the study were typical antipsychotics (haloperidol, perphenazine) while only 1 trial studied an SGA, risperidone. Among the five studies comparing SSRIs with placebo, two of them reported a significant benefit for Citalopram against AD+P ^22, 24^. Meanwhile, no significant difference was reported between the efficacy of SSRIs and risperidone. Therefore, testing more antidepressants, especially other classes of antidepressants such as serotonin-noradrenaline reuptake inhibitors, tricyclic antidepressants, and monoamine oxidase inhibitors, may be worthwhile to provide a better understanding of the impact of antidepressants on AD+P.

While antipsychotics and antidepressants both have shown beneficial effects against AD+P, they have different targets which in turn can modulate different biological pathways. Thus, the combination of these drugs can potentially provide multiple advantages like enhanced efficacy, decreased dosage with an equal or increased level of efficacy, and delayed development of drug resistance ^33^. Due to the excessive time and cost it takes to clinically test the drug combination effects, exhaustive computational methods can be used to predict drug synergy. By integrating information from drugs and diseases we can obtain a comprehensive picture of the potential synergetic effects of these drug combinations. The goal of our study is to further identify key combinations of antipsychotics and antidepressants that possess potential synergetic effects against AD+P with the help of our state-of-the-art quantitative systems pharmacology approaches.

## 2. Methods and Material

### 2.1 Dataset collection

To explore the beneficial effect of combination therapy, we examined the data from January 2004 to October 2019 from the Neptune system at the University of Pittsburgh Medical Center (UPMC), which manages the use of patient EMRs from the UPMC health system for research purposes (rio.pitt.edu/services)^34^. The database includes demographic information, diagnoses, encounters, medication prescriptions, prescription fill history, and laboratory tests. AD patients were identified using ICD9/10 codes (331.0, G30.0, G30.1, G30.9) and the onset of psychosis were defined by ICD9/10 codes (780.1, F06.0, R44.2, R44.1, R44.3, R44.0, 298.8, F22, F23, F28, F29, 293.82, 298.9, 290.11, 293, 290.3) based on the suggestions from UPMC clinicians.

### 2.2 Survival analysis

We included patients who met the following inclusion criteria: 1) Patient had an AD diagnosis; 2) Patients did not take antidepressants nor antipsychotics one year prior to the diagnosis of AD. Nine comorbidities, including MDD (major depressive disorder), stroke, COPD (chronic obstructive pulmonary disease), ASCVD (atherosclerotic cardiovascular disease), T2DM (type 2 diabetes), HTN (hypertension), CKD (chronic kidney disease), HF (heart failure) and cancer, were considered as confounders in our survival analysis (ICD codes listed in supplementary material) ^35, 36^. The time origin for each patient in the survival analysis is the first AD diagnosis date and time to all-cause death is the outcome. Patients are marked with the above comorbidities if they were diagnosed before the AD diagnosis. Only medications that are prescribed to the same patients more than 2 times with more than 30 days apart were considered to eliminate short-term usage during hospitalization. The records of patients up to 5 years after the AD diagnosis were used in the analysis.

Time to all-cause death was constructed as the time between the first date of AD diagnosis and death. Patients who were alive by the end of 5 years since AD diagnosis were censored. Survival analysis was performed to evaluate the association between medications and mortality. To accommodate the change of drug usage during the follow-up, we fitted a time-dependent Cox’s proportional hazards model^37, 38^ with antipsychotic drug effect (yes or no) and antidepressant drug effect (yes or no) as time dependent covariates. Specifically, the 5-year follow-up period was divided into 60 months, and we assumed the drug effect from one prescription will last 2 months which covers 2 intervals in our study. As shown in **Figure 1**, if a patient had an antipsychotics prescription in the first month, we consider the patient under drug effect for that month and the month after. If a patient was prescribed both antipsychotics and antidepressants (boxed in **Figure 1**), we consider the patient under combinational therapy. The drug effect on one patient may change over time between four statuses: no drug, antipsychotics only, antidepressants only, and the combination. Baseline demographics and comorbidities were also included in the model. Contrasts between different drug groups were performed with hazard ratios and p values reported. The data were analyzed using both R (version 4.1.0) and python (version 3.7.12) packages.

**Figure 1.**
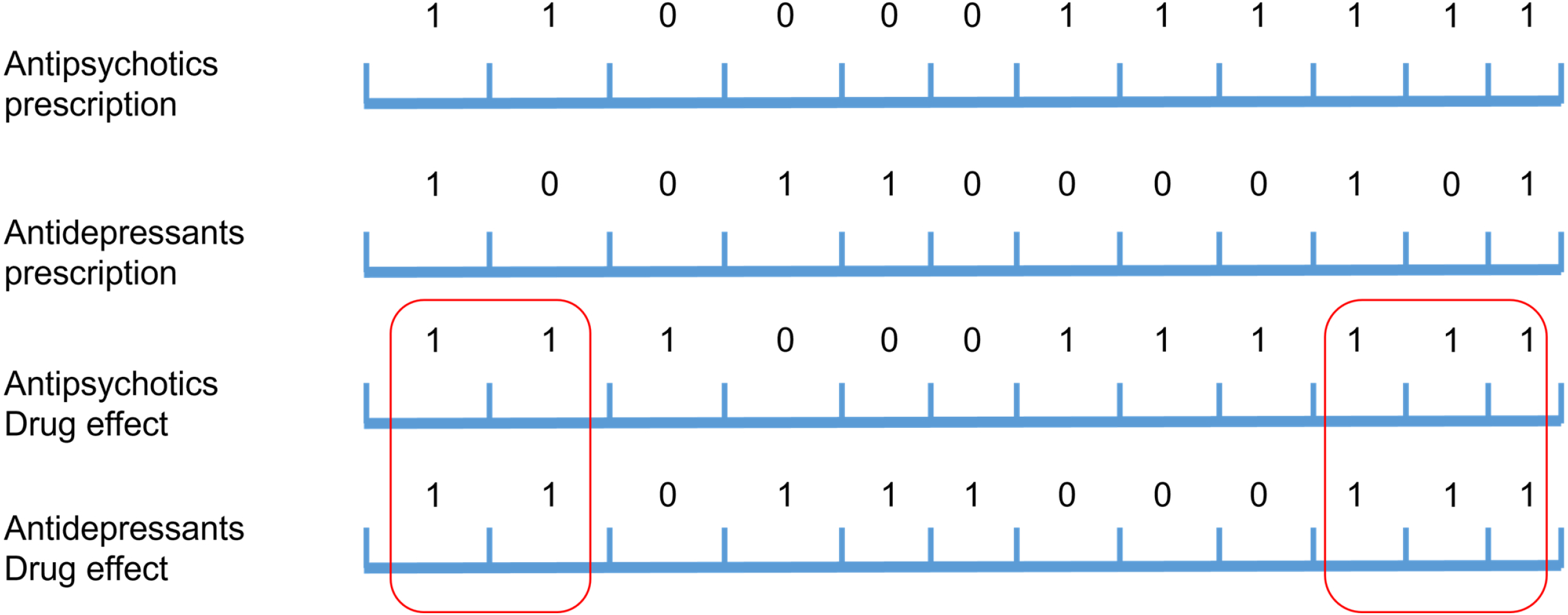
Schematic diagram for identifying drug usage status of subjects. In the upper two rows of Figure 1, the sections with 1 indicate that there are antipsychotics/antidepressants prescriptions in that month while 0 means there are no prescriptions for the two kinds of medications. In the lower rows, the sections under 1 are extended for 1 month to reflect the drug effect thus these markers show the time that the subject was under drug effect.

### 2.3 Prediction of synergetic effect among antipsychotic-antidepressant pairs

Since the analysis conducted are a mixed effect of any combination of antipsychotics and antidepressants, we further examined if certain antipsychotic-antidepressant pairs may possess higher synergetic effect in managing psychotic symptoms in AD. We first attempted this evaluation by using EMR data. However, because of the large number of antipsychotics and antidepressants used clinically, the appearances of specific drug pairs are limited (most co-administrated pair: Quetiapine and Sertraline, 408 times. More data in supplementary material **ST1**), and their overlap times are too short to produce enough statistical power. Therefore, we turned to systems pharmacology approaches to identify antipsychotic-antidepressant pairs that are most suited in treating AD+P.

For the systems pharmacology study, the postsynaptic density (PSD) proteome was used to build the AD+P network. This proteomic signature was generated by Dr. Sweet’s team^39^. Information about antipsychotics, antidepressants and their targets were extracted from DrugBank (https://www.drugbank.ca/)^40^. The pharmacological action label of a drug provides information about whether binding to a target contributes to the pharmacological effects. PPI data was collected from STRING (https://string-db.org/)^41^. The PPI networks were constructed and analyzed using the python package networkx (https://networkx.github.io/)^42^. The interaction network was shown in the molecular action view with medium confidence level (> 0.4) ^43^. AD+P-related proteins were joined with targets of antipsychotics and antidepressants to construct the disease-target network. In addition, we also included the proteins bridging proteins from disease module and proteins from the target module in our disease-targets networks. Gene signature data were used to calculate the proximity score for drugs and AD+P. The post-treatment gene signature data were obtained from the LINCS L1000 database^44^.

To predict potential drug combinations for AD+P, we adopted the methods from Chen, S., et al. ^45^ and modified them by incorporating differentially expressed genes (DEGs) after drug treatment to minimize the bias caused by module sizes. Based on previous studies, for a drug pair to have a therapeutic effect on a disease, both target modules (green and yellow circles in **figure 2**) of the two drugs must overlap with the disease module (pink circle in **figure 2**) ^45^. In addition, the two target modules need to be overlapped with the disease module independently to form a complementary exposure to have synergetic effects with each other as shown in **Figure 2**. To be specific, the targets of the two drugs both need to be overlapped with the disease module in the PPI network, but these two target modules can’t overlap ^45^. Therefore, two network approaches are applied to predict the possible drug combinations for AD+P: (1) network-based separation between targets of two drugs ^46^; (2) gene signature-based proximity between the disease (AD+P) module and the target modules of the two drugs ^47^.

**Figure 2.**
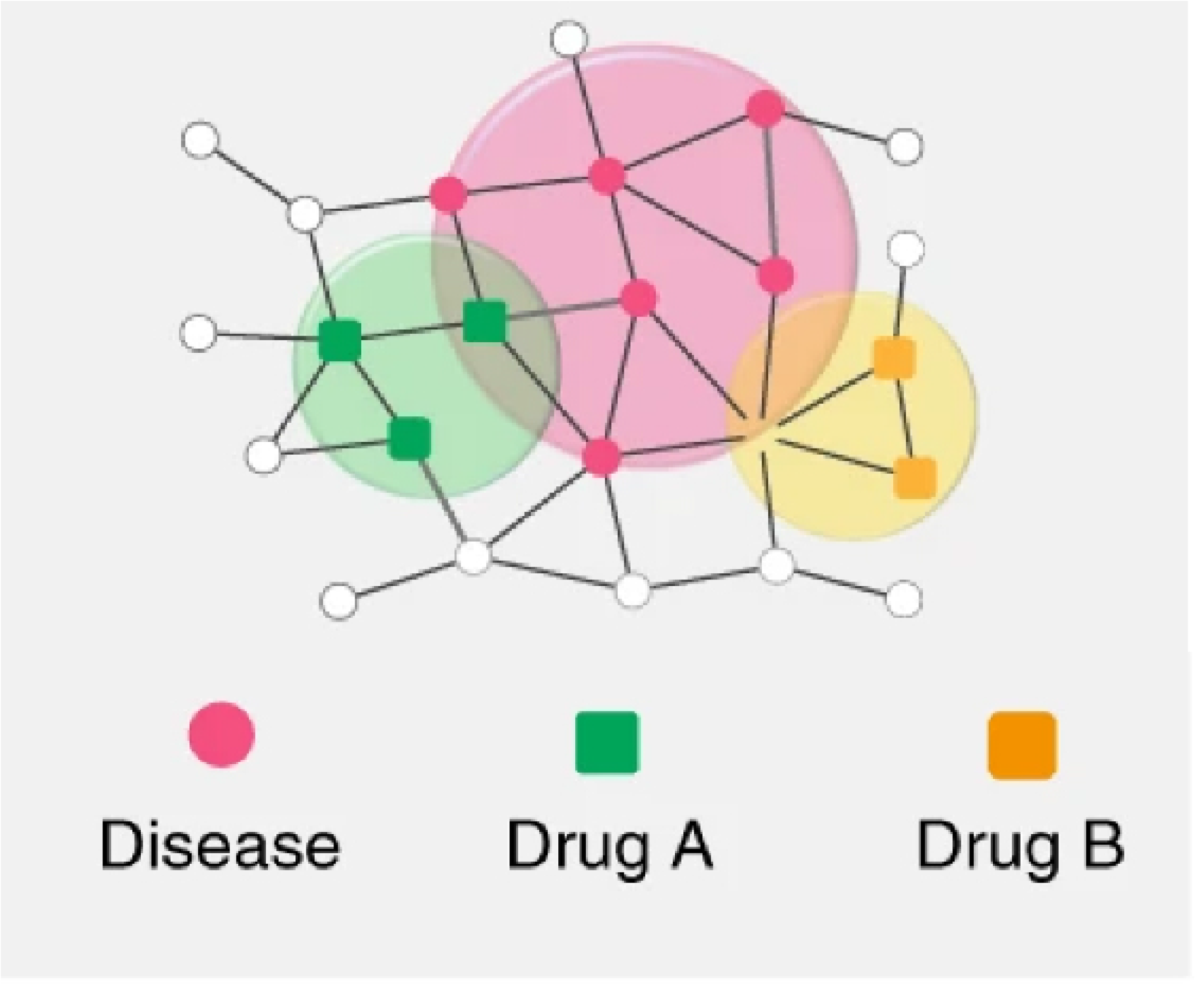
Schematic diagram for the network-based complementary exposure relationship between two drug–target modules and one disease module on a drug–drug–disease combination (adopted from Guney, E.; Menche, J.; Vidal, M.; Barábasi, A.-L., Network-based in silico drug efficacy screening. Nature communications 2016, 7 (1), 1-13. ^47^). Big red circle: AD+P modules composed of AD+P related proteins/genes (small red rounds). Green and yellow circle: Drug modules composed of drug targets.

#### Separation evaluation

The separation score (S_AB_) of drug modules A and B are calculated for all possible combinations between antipsychotics and antidepressants. The separation score (S_AB_) of drug modules A and B can be calculated as:

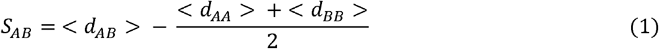

where <d_AA_>, <d_BB_> and <d_AB_>, are the mean shortest distance for genes within each module. It compares the mean shortest distance between the targets of each drug. For better understanding, if S_AB_ < 0, it means that the targets in the two drug modules are in the same network neighborhood which is not separated; if S_AB *≥*_ 0, it means that the two drug modules are topologically separated from each other. We filter the combinations based on their ability to achieve complementary exposure with the AD+P module.

#### Proximity evaluation

Level 5 LINCS L1000 data, a collection of gene expression profiles for thousands of perturbagens at a variety of time points, doses, and cell lines, were downloaded from the GEO database (accession numbers: GSE70138 and GSE92742). Gene expression profiles were included only if they are tested on a cell line of central nervous systems and their dose should be beyond 1 uM. To identify genes that are significantly differentially expressed in the data, their Z scores from multiple tests were averaged and if their |Z|>1, the genes are considered as significant for a drug^48^.

The association between the drug and AD+P was quantitatively evaluated with Signed Jaccard Score^49^. The index ranges from +1 to *−*1, where +1 and *−*1 indicate a same pattern and an inverse pattern of two identical gene sets, respectively. Zero indicates that the two sets have no overlap, or the positive and negative correlations cancel out.

## 3. Results

After applying all inclusion criteria to our dataset, 10,206 unique AD patients were included, and their baseline characteristics are shown in **supplementary material (ST2)**.

### 3.1 Use of antipsychotics in AD patients is associated with increased mortality

cPrior literature has reported that the use of antipsychotics is associated with increased mortality in AD patients^50-52^; we therefore first sought to replicate this finding to validate the integrity of our methodological approach. The time varying Cox model was conducted with our data to test the significance.

**Table 1** Multivariate Cox regression analyses of association between antipsychotics and all-cause mortality in AD patients As indicated in **Table 1**, antipsychotic usage is significantly associated with increased mortality in AD patients (HR=2.47, p<0.001). The results showed in this table are in accordance with current literature which suggest that our data and methods are valid for further analysis.

### 3.2 Survival analysis revealed significant beneficial effect combining antidepressants and antipsychotics in AD patients

After knowing that antipsychotics may increase mortality in AD patients while antidepressants may decrease mortality^53^, the effect of a combination therapy comes into play. We would like to examine the protective effects of adding antidepressants to the existing antipsychotics therapy. We performed another survival analysis to examine three mutually exclusive medication use groups: antipsychotics only, antidepressants only and combination.

The results are shown in **Table 2** and the combination group is the reference group in this model. Based on the results from **Table 2**, the combination group showed a significant beneficial effect relative to antipsychotics only group (HR=0.654, p=0.012), which means that combining antidepressants with antipsychotic treatment was associated with significantly protective effects in AD patients, reducing mortality. In addition, marginal significant difference was observed between no drug group and combination group (HR=1.294, p=0.056), which means that by using combination therapy, the increase in mortality due to using antipsychotics was mitigated to some extent in these patients.

**Table 2** Multivariate Cox regression analyses of association among treatments and all-cause mortality in AD patients

Based on our findings shown in **Table 1** and **Table 2**, we can conclude that by combining antipsychotics and antidepressants, we can significantly mitigate the increase in mortality associated with antipsychotics. For more direct comparison between different treatment groups, a table with pair-wise comparison among groups is included in supplementary table 2 (**ST3**).

In addition to these results, we were interested to see if the effect of the treatments will change over time. Therefore, we conducted 6 analyses with follow-up times ranging from 1 to 6 years. We compared the effects of three different treatments (antipsychotics only, antidepressants only, no drug) to the drug combination group, their effects showed moderate fluctuation within the first 3 years and stabilized after 4 years (**SF1**). In comparison to co-administration of antidepressants and antipsychotics, the antidepressants only group showed a consistent lower mortality throughout the 6 years. While the antipsychotics only group had marginally increased mortality compared to the combination group at the first 3 years, subsequent worse outcomes were clearly evident in 4, 5 and 6 years of follow-up. Finally, the combination group showed comparable effects with the patients with no drug treatment throughout the 6 years period and demonstrated almost significant beneficial effects in 4, 5 and 6 years of the follow-up. Our results are attached in supplementary materials (**ST4**).

### 3.2 Systems pharmacology studies provided possible explanations for the synergetic effects and proposed drug pairs may achieve optimized efficacy

In total, 21 antipsychotics and 17 antidepressants commonly used in the clinic are included in our study along with 75 targets for antipsychotics and 32 targets for antidepressants. The PPI network was built with 240 AD+P proteins, targets for antipsychotics and antipsychotics. A PPI network with 321 nodes and 1,363 edges was generated. A total of 357 pairs of antipsychotics and antidepressants are evaluated in the network and their separation scores are calculated as shown in **supplementary figure (SF2)**.

We found that some antidepressants showed great separation (Vortioxetine, Vilazodone, Mirtazapine, Maprotiline), and most drugs pairs showed a separation score above 0 (blue). This suggests an existing difference in the mechanism which can be the key condition to the synergetic effect in the combinational therapy.

To evaluate the proximity between AD+P and medications, 148 and 78 eligible expression profiles for antipsychotics and antidepressants were collected based on the inclusion criteria described earlier. Signed Jaccard scores were calculated between them and AD+P protein expressions.

Post-treatment gene expression data for 16 antipsychotics and 13 antidepressants were exacted, and their Signed Jaccard scores were calculated accordingly and are shown in the tables below.

**Table 3** Signed Jaccard scores of antipsychotics and antidepressants with AD+P

To comprehensively evaluate the potential for these drug combinations, separation scores and proximity scores were normalized to a [-1, 1] interval and combined. A combined score was calculated for every drug pair by subtracting two proximity scores of the drugs from their separation score. The combined scores for drug pairs are shown in the heatmap below (**Figure 5**).

As shown in **Figure 3**, most drug pairs showed a combined score around 0 which indicate that the synergistic effect between antipsychotics and antidepressants may not be easily achieved within the two drug categories. However, some drugs showed promising results through the panel like Aripiprazole and Thioridazine. For drug combinations that may possess beneficial synergistic effect, they must meet the following criteria: 1) The separation score between two drugs > 0; 2) The proximity scores of the two drugs <0. **Table 4** summarized the drug combinations that met the criteria.

**Figure 3.**
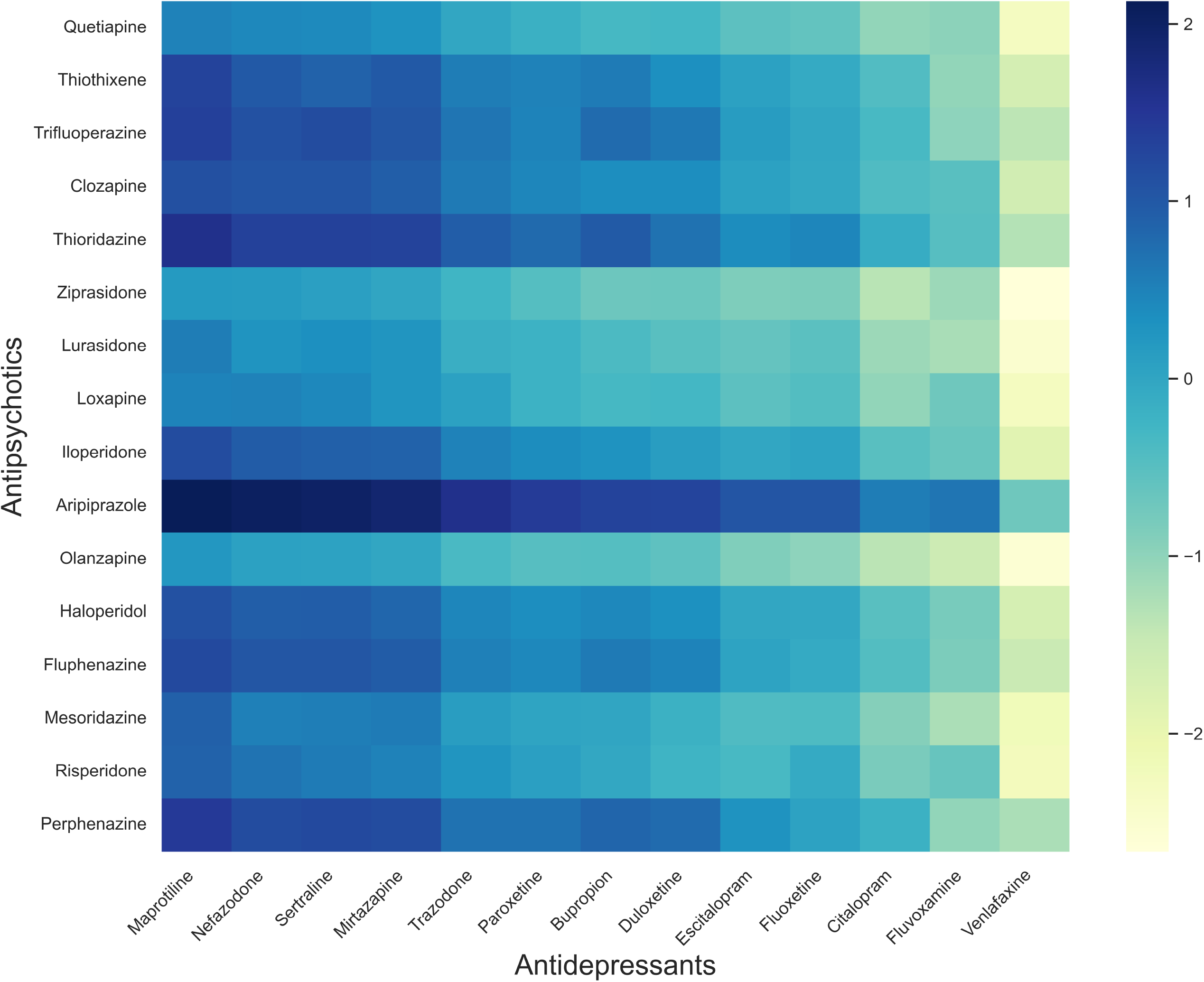
Combined scores for antipsychotics and antidepressants combinations. Though most drug pairs showed a combined score around 0, some drugs do show promising scores across the board including Aripiprazole and Thioridazine.

**Table 4** Antidepressants and antipsychotics combinations with highest combined scores

Among the nominated drug combinations, Aripiprazole is the only SGAs and is also the most recommended treatment options for AD+P. As a matter of fact, the combination of Aripiprazole and Sertraline has already been tested in clinical trials and showed superior efficacy in treating major depressive disorders (MDD) with no reports of safety concerns^54^. However, none of these drug pairs have been tested for AD+P in clinical trials. As a matter of fact, no combination drug therapy has ever been tested for AD+P at present.

## 4. Discussion and conclusion

In this study, we applied a combination of different state-of-the-art systems pharmacology techniques to explore the potential synergetic effect of combining antipsychotics and antidepressants in treating AD+P. This study incorporated different data types including clinical EMR data, protein expressions, post-treatment gene expressions and protein-protein interaction networks. Our results indicate that the combination of antipsychotics and antidepressants may be a safer and more efficient treatment option for AD+P and provided informative insights for drug pairing choices for future therapies.

When dealing with real-world data, like EMRs, there is always a challenge that the compliance of patients presented in the EMR will not be as ideal as we get from a carefully performed clinical trial. In this study, by analyzing real-world EMR data through Cox model with time-dependent covariates, we were able to accommodate the complex usage patterns and allowed the maximum utilization of the data. The beneficial effect of antidepressants in AD patients were reported by multiple studies^55-58^, though its mechanisms remain unclear. We also found a strong signal in our results (**Table 3** and **Figure 3**) which further substantiated our claim that antidepressants may aid in reducing mortality in AD patients. Our finding provided the fundamental support necessary for our hypothesis that by combining antipsychotics and antidepressants, we can decrease the severe side effect that is constraining the use of antipsychotics in AD therapy.

In order to provide mechanistic support for our observations and identify the most potent drug combinations for treating AD+P, we took advantage of multiple categories of data including PPI network, post-treatment gene expression profiles to quantitatively evaluate the potential synergistic effect of antipsychotics and antidepressants. Our analysis yielded several pairs of drugs that may possess better synergistic effects in treating AD+P. As shown in **Table 6**, four antipsychotics: Aripiprazole, Thioridazine, were reported, and seven antidepressants: Sertraline, Maprotiline, Nefazodone, Mirtazapine, Trazodone, Paroxetine and Bupropion were mentioned. Between the 2 antipsychotics, Thioridazine is first-generation antipsychotic and it was withdrawn worldwide in 2005 due to its association with cardiac arrythmias^59^. For Aripiprazole, it has been reported with significant better efficacy in AD patients against psychological symptoms^60^ which further consolidate our conclusions. For the 7 antidepressants, Nefazodone is discontinued on 2004 because of its association with drug-induced hepatic injuries^61^. For the rest 6 antidepressants, they belong to 4 classes. Sertraline, Trazodone, Paroxetine are selective serotonin reuptake inhibitors (SSRI) which is the most used antidepressant class^62^. Maprotiline is a tetracyclic antidepressant with similar pharmacological properties to tricyclic antidepressants (TCAs), it can inhibit neuronal norepinephrine reuptake, possesses some anticholinergic activity, and does not affect monoamine oxidase activity^63^. Mirtazapine is a tetracyclic *piperazino-azepine* antidepressant with its effect can be observed as early as 1 week after beginning therapy^64^, it has also been reported to be efficacious in the off-label management of various other conditions. It may improve the symptoms of neurological disorders, reverse weight loss caused by medical conditions, improve sleep, and prevent nausea and vomiting after surgery^65^. Bupropion is a norepinephrine/dopamine-reuptake inhibitor (NDRI) antidepressant, and it is a unique option for the treatment of MDD as it lacks any clinically relevant serotonergic effects, typical of other mood medications, or any effects on histamine or adrenaline receptors^66, 67^. As for combination therapy consisting of antipsychotics and antidepressants, though they were never tested specifically for AD or AD+P, multiple studies have tested their safety and efficacy profile against other disorders. For example, a meta-analysis consisting of eight randomized, placebo-controlled studies reported that antidepressant-antipsychotic cotreatment was superior to monotherapy with either drug class in the acute treatment of psychotic depression^68^ and another study reported that adding SGAs to antidepressants yielded highly significant superiority in treating MDD, too^69^. In that study, Aripiprazole, Olanzapine, Risperidone, and Ziprasidone were found to be more effective than other SGAs^69^.

The results of our study aims to provide a comprehensive and quantitative overview of the underlying relationship among antipsychotics, antidepressants, and AD+P. Our results supported the efficacy of antipsychotics and suggested the most promising antidepressants such as Sertraline and Maprotiline, can be added as supplementary treatment. In addition, since they are all marketed drugs and some of their combinations are already tested by clinical trials for other indications, safety profile will not be a major concern when proposing their long-term usage as an alternative treatment option for AD patients.

## Data Availability

All data produced in the present study are available upon reasonable request to the authors

## 5. Author Contribution

P.F. wrote the manuscript; P.F., Z.L., Y.D. and L.W. designed the research; P.F. and Z.L. performed the research; P.F. and Z.L. analyzed the data; J.K., J.S., J.K. and R.A.S. contributed new reagents/analytical tools.

## 6. Funding

This study is supported by the following National Institutes of Health grants:

S10 OD028483/OD/NIH HHS/United States

P50 AG005133/AG/NIA NIH HHS/United States

R01 AG027224/AG/NIA NIH HHS/United States

R01 MH116046/MH/NIMH NIH HHS/United States

P30 AG066468/AG/NIA NIH HHS/United States

UL1 TR001857/TR/NCATS NIH HHS/United States

## 7. Study Highlights

- What is the current knowledge on the topic? Current treatments to AD+P are restricted by their modest efficacy and poor safety profiles. Better treatment options are in urgent need.
- What question did this study address? Will the combination of antipsychotics and antidepressants be a better treatment option? Which antipsychotics and antidepressants are the best pair?
- What does this study add to our knowledge? This study proved that the combination of antipsychotics and antidepressants showed superiority over antipsychotics monotherapy and proposed several drug pairs that may possess higher efficacy.
- How might this change drug discovery, development, and/or therapeutics?

Our results supported the efficacy of antipsychotics and suggested the most promising antidepressants such as Sertraline and Maprotiline, can be added as supplementary treatment. In addition, since they are all marketed drugs and some of their combinations are already tested by clinical trials for other indications, safety profile will not be a major concern when proposing their long-term usage as an alternative treatment option for AD patients.

## References

1. Murray, P. S.; Kumar, S.; Demichele-Sweet, M. A.; Sweet, R. A., Psychosis in Alzheimer’s disease. Biological psychiatry 2014, 75 (7), 542–52.

2. Ropacki, S. A.; Jeste, D. V., Epidemiology of and risk factors for psychosis of Alzheimer’s disease: a review of 55 studies published from 1990 to 2003. The American journal of psychiatry 2005, 162 (11), 2022–30.

3. Gilley, D. W.; Wilson, R. S.; Beckett, L. A.; Evans, D. A., Psychotic symptoms and physically aggressive behavior in Alzheimer’s disease. Journal of the American Geriatrics Society 1997, 45 (9), 1074–9.

4. Sweet, R. A.; Pollock, B. G.; Sukonick, D. L.; Mulsant, B. H.; Rosen, J.; Klunk, W. E.; Kastango, K. B.; DeKosky, S. T.; Ferrell, R. E., The 5-HTTPR polymorphism confers liability to a combined phenotype of psychotic and aggressive behavior in Alzheimer disease. International psychogeriatrics 2001, 13 (4), 401–9.

5. Lyketsos, C. G.; Sheppard, J. M.; Steinberg, M.; Tschanz, J. A.; Norton, M. C.; Steffens, D. C.; Breitner, J. C., Neuropsychiatric disturbance in Alzheimer’s disease clusters into three groups: the Cache County study. International journal of geriatric psychiatry 2001, 16 (11), 1043–53.

6. Sweet, R. A.; Bennett, D. A.; Graff-Radford, N. R.; Mayeux, R., Assessment and familial aggregation of psychosis in Alzheimer’s disease from the National Institute on Aging Late Onset Alzheimer’s Disease Family Study. Brain : a journal of neurology 2010, 133 (Pt 4), 1155–62.

7. Wilson, R. S.; Tang, Y.; Aggarwal, N. T.; Gilley, D. W.; McCann, J. J.; Bienias, J. L.; Evans, D. A., Hallucinations, cognitive decline, and death in Alzheimer’s disease. Neuroepidemiology 2006, 26 (2), 68–75.

8. Scarmeas, N.; Brandt, J.; Albert, M.; Hadjigeorgiou, G.; Papadimitriou, A.; Dubois, B.; Sarazin, M.; Devanand, D.; Honig, L.; Marder, K.; Bell, K.; Wegesin, D.; Blacker, D.; Stern, Y., Delusions and hallucinations are associated with worse outcome in Alzheimer disease. Archives of neurology 2005, 62 (10), 1601–8.

9. Kaufer, D. I.; Cummings, J. L.; Christine, D.; Bray, T.; Castellon, S.; Masterman, D.; MacMillan, A.; Ketchel, P.; DeKosky, S. T., Assessing the impact of neuropsychiatric symptoms in Alzheimer’s disease: the Neuropsychiatric Inventory Caregiver Distress Scale. Journal of the American Geriatrics Society 1998, 46 (2), 210–5.

10. Madhusoodanan, S.; Shah, P., Management of psychosis in patients with Alzheimer’s disease: focus on aripiprazole. Clin Interv Aging 2008, 3 (3), 491–501.

11. Alexopoulos, G. S.; Streim, J.; Carpenter, D.; Docherty, J. P., Using antipsychotic agents in older patients. The Journal of clinical psychiatry 2004, 65 Suppl 2, 5–99; discussion 100-102; quiz 103-4.

12. Burke, A. D.; Burke, W. J., Antipsychotics FOR patients WITH dementia: The road less traveled: Second-generation agents have an important but limited role in treating behavioral and psychological symptoms. Current Psychiatry 2018, 17 (10), 26–36.

13. Kindermann, S. S.; Dolder, C. R.; Bailey, A.; Katz, I. R.; Jeste, D. V., Pharmacological treatment of psychosis and agitation in elderly patients with dementia: four decades of experience. Drugs & aging 2002, 19 (4), 257–76.

14. Masand, P. S., Side effects of antipsychotics in the elderly. The Journal of clinical psychiatry 2000, 61 Suppl 8, 43–9; discussion 50-1.

15. Dorsey, E. R.; Rabbani, A.; Gallagher, S. A.; Conti, R. M.; Alexander, G. C., Impact of FDA black box advisory on antipsychotic medication use. Archives of internal medicine 2010, 170 (1), 96–103.

16. Tampi, R. R.; Tampi, D. J.; Balachandran, S.; Srinivasan, S., Antipsychotic use in dementia: a systematic review of benefits and risks from meta-analyses. Ther Adv Chronic Dis 2016, 7 (5), 229–245.

17. Gallagher, D.; Herrmann, N., Agitation and aggression in Alzheimer’s disease: an update on pharmacological and psychosocial approaches to care. Neurodegenerative disease management 2015, 5 (1), 75–83.

18. Madhusoodanan, S.; Ting, M. B., Pharmacological management of behavioral symptoms associated with dementia. World J Psychiatry 2014, 4 (4), 72–79.

19. Fan, P.; Kofler, J.; Ding, Y.; Marks, M.; Sweet, R. A.; Wang, L., Efficacy difference of antipsychotics in Alzheimer’s disease and schizophrenia: explained with network efficiency and pathway analysis methods. Briefings in Bioinformatics 2022.

20. Sweet, R. A.; Bennett, D. A.; Graff-Radford, N. R.; Mayeux, R.; Group*, t. N. I. o. A. L.-O. A. s. D. F. S., Assessment and familial aggregation of psychosis in Alzheimer’s disease from the National Institute on Aging Late Onset Alzheimer’s Disease Family Study. Brain : a journal of neurology 2010, 133 (4), 1155–1162.

21. DeMichele-Sweet, M. A. A.; Klei, L.; Creese, B.; Harwood, J. C.; Weamer, E. A.; McClain, L.; Sims, R.; Hernandez, I.; Moreno-Grau, S.; Tárraga, L.; Boada, M.; Alarcón-Martín, E.; Valero, S.; Liu, Y.; Hooli, B.; Aarsland, D.; Selbaek, G.; Bergh, S.; Rongve, A.; Saltvedt, I.; Skjellegrind, H. K.; Engdahl, B.; Stordal, E.; Andreassen, O. A.; Djurovic, S.; Athanasiu, L.; Seripa, D.; Borroni, B.; Albani, D.; Forloni, G.; Mecocci, P.; Serretti, A.; De Ronchi, D.; Politis, A.; Williams, J.; Mayeux, R.; Foroud, T.; Ruiz, A.; Ballard, C.; Holmans, P.; Lopez, O. L.; Kamboh, M. I.; Devlin, B.; Sweet, R. A.; Nia-Load Family Based Study Consortium, A. s. D. G. C., Genome-wide association identifies the first risk loci for psychosis in Alzheimer disease. Molecular Psychiatry 2021.

22. Auchus, A. P.; Bissey-Black, C., Pilot study of haloperidol, fluoxetine, and placebo for agitation in Alzheimer’s disease. The Journal of neuropsychiatry and clinical neurosciences 1997, 9 (4), 591–593.

23. McRae, T., Donepezil and sertraline for the management of behavioral symptoms in patients with Alzheimer’s disease. Neurology 2000, 54, A416.

24. Finkel, S. I.; Mintzer, J. E.; Dysken, M.; Krishnan, K.; Burt, T.; McRae, T., A randomized, placebo - controlled study of the efficacy and safety of sertraline in the treatment of the behavioral manifestations of Alzheimer’s disease in outpatients treated with donepezil. International journal of geriatric psychiatry 2004, 19 (1), 9–18.

25. Finkel, S.; Mintzer, J.; Burt, T.; McRae, T., Sertraline augmentation reduces behavioral symptoms in outpatients with Alzheimer’s disease treated with donepezil. European Neuropsychopharmacology 2002, (12), 374.

26. Gaber, S.; Ronzoli, S.; Bruno, A.; Biagi, A., Sertraline versus small doses of haloperidol in the treatment of agitated behavior in patients with dementia. Archives of Gerontology and Geriatrics 2001, 33, 159–162.

27. Elgen, K.; Gottfries, C.-G.; Nyth, A. L., Effekt av citalopram på emosjonelle forstyrrelser hos pasienter med Alzheimer demens. Nordisk Psykiatrisk Tidsskrift 1991, 45 (sup23), 23–27.

28. Nyth, A. L.; Gottfries, C. G., The clinical efficacy of citalopram in treatment of emotional disturbances in dementia disorders A Nordic multicentre study. The British Journal of Psychiatry 1990, 157 (6), 894–901.

29. Olafsson, K.; Jørgensen, S.; Jensen, H.; Bille, A.; Arup, P.; Andersen, J., Fluvoxamine in the treatment of demented elderly patients: a double-blind, placebo-controlled study. Acta Psychiatrica Scandinavica 1992, 85 (6), 453–456.

30. Pollock, B.; Rosen, J.; Mulsant, B. In Placebo-controlled comparison of citalopram versus perphenazine for acute treatment of severe behavioral disturbances associated with dementia, Annual Meeting of the American Association for Geriatric Psychiatry, 2001.

31. Pollock, B. G.; Mulsant, B. H.; Rosen, J.; Sweet, R. A.; Mazumdar, S.; Bharucha, A.; Marin, R.; Jacob, N.; Huber, K. A.; Kastango, K. B., Comparison of citalopram, perphenazine, and placebo for the acute treatment of psychosis and behavioral disturbances in hospitalized, demented patients. American Journal of Psychiatry 2002, 159 (3), 460–465.

32. Pollock, B. G.; Mulsant, B. H.; Rosen, J.; Mazumdar, S.; Blakesley, R. E.; Houck, P. R.; Huber, K. A., A double-blind comparison of citalopram and risperidone for the treatment of behavioral and psychotic symptoms associated with dementia. The American Journal of Geriatric Psychiatry 2007, 15 (11), 942–952.

33. Nelson, H. S., Advair: combination treatment with fluticasone propionate/salmeterol in the treatment of asthma. Journal of allergy and clinical immunology 2001, 107 (2), 397–416.

34. Visweswaran, S.; McLay, B.; Cappella, N.; Morris, M.; Milnes, J. T.; Reis, S. E.; Silverstein, J. C.; Becich, M. J., An atomic approach to the design and implementation of a research data warehouse. Journal of the American Medical Informatics Association 2022, 29 (4), 601–608.

35. Fitzpatrick, A. L.; Kuller, L. H.; Lopez, O. L.; Kawas, C. H.; Jagust, W., Survival following dementia onset: Alzheimer’s disease and vascular dementia. Journal of the Neurological Sciences 2005, 229–230, 43–49.

36. Go, S. M.; Lee, K. S.; Seo, S. W.; Chin, J.; Kang, S. J.; Moon, S. Y.; Na, D. L.; Cheong, H. K., Survival of Alzheimer’s Disease Patients in Korea. Dementia and Geriatric Cognitive Disorders 2013, 35 (3-4), 219–228.

37. Tian, L.; Zucker, D.; Wei, L. J. J. o. t. A. s. A., On the Cox model with time-varying regression coefficients. 2005, 100 (469), 172–183.

38. Therneau, T.; Crowson, C.; Atkinson, E. J. S. V., Using time dependent covariates and time dependent coefficients in the cox model. 2017, 2 (3), 1–25.

39. Krivinko, J.; DeChellis-Marks, M.; Zeng, L.; Fan, P.; Lopez, O.; Ding, Y.; Wang, L.; Kofler, J.; MacDonald, M.; Sweet, R., Targeting the Post-Synaptic Proteome in Alzheimer Disease with Psychosis. Research Square: 2022.

40. Wishart, D. S.; Feunang, Y. D.; Guo, A. C.; Lo, E. J.; Marcu, A.; Grant, J. R.; Sajed, T.; Johnson, D.; Li, C.; Sayeeda, Z.; Assempour, N.; Iynkkaran, I.; Liu, Y.; Maciejewski, A.; Gale, N.; Wilson, A.; Chin, L.; Cummings, R.; Le, D.; Pon, A.; Knox, C.; Wilson, M., DrugBank 5.0: a major update to the DrugBank database for 2018. Nucleic Acids Res 2018, 46 (D1), D1074–d1082.

41. Jensen, L. J.; Kuhn, M.; Stark, M.; Chaffron, S.; Creevey, C.; Muller, J.; Doerks, T.; Julien, P.; Roth, A.; Simonovic, M.; Bork, P.; von Mering, C., STRING 8--a global view on proteins and their functional interactions in 630 organisms. Nucleic Acids Res 2009, 37 (Database issue), D412–6.

42. Aric A. Hagberg, D. A. S., Pieter J. Swart Exploring Network Structure, Dynamics, and Function using NetworkX, Proceedings of the 7th Python in Science Conference, 2008.

43. Franceschini, A.; Szklarczyk, D.; Frankild, S.; Kuhn, M.; Simonovic, M.; Roth, A.; Lin, J.; Minguez, P.; Bork, P.; Von Mering, C., STRING v9. 1: protein-protein interaction networks, with increased coverage and integration. Nucleic acids research 2012, 41 (D1), D808–D815.

44. Liu, C.; Su, J.; Yang, F.; Wei, K.; Ma, J.; Zhou, X., Compound signature detection on LINCS L1000 big data. Molecular BioSystems 2015, 11 (3), 714–722.

45. Cheng, F.; Kovács, I. A.; Barabási, A.-L., Network-based prediction of drug combinations. Nature Communications 2019, 10 (1), 1197.

46. Menche, J.; Sharma, A.; Kitsak, M.; Ghiassian, S. D.; Vidal, M.; Loscalzo, J.; Barabási, A.-L., Uncovering disease-disease relationships through the incomplete interactome. Science 2015, 347 (6224).

47. Guney, E.; Menche, J.; Vidal, M.; Barábasi, A.-L., Network-based in silico drug efficacy screening. Nature communications 2016, 7 (1), 1–13.

48. Stathias, V.; Jermakowicz, A. M.; Maloof, M. E.; Forlin, M.; Walters, W.; Suter, R. K.; Durante, M. A.; Williams, S. L.; Harbour, J. W.; Volmar, C.-H.; Lyons, N. J.; Wahlestedt, C.; Graham, R. M.; Ivan, M. E.; Komotar, R. J.; Sarkaria, J. N.; Subramanian, A.; Golub, T. R.; Schürer, S. C.; Ayad, N. G., Drug and disease signature integration identifies synergistic combinations in glioblastoma. Nature Communications 2018, 9 (1), 5315.

49. Qi, X.; Shen, M.; Fan, P.; Guo, X.; Wang, T.; Feng, N.; Zhang, M.; Sweet, R. A.; Kirisci, L.; Wang, L., The Performance of Gene Expression Signature-Guided Drug–Disease Association in Different Categories of Drugs and Diseases. 2020, 25 (12), 2776.

50. Nielsen, R.; Lolk, A.; Valentin, J.; Andersen, K. J. A. P. S., Cumulative dosages of antipsychotic drugs are associated with increased mortality rate in patients with Alzheimer’s dementia. 2016, 134 (4), 314–320.

51. Schwertner, E.; Secnik, J.; Garcia-Ptacek, S.; Johansson, B.; Nagga, K.; Eriksdotter, M.; Winblad, B.; Religa, D. J. J. o. t. A. M. D. A., Antipsychotic treatment associated with increased mortality risk in patients with dementia. A registry-based observational cohort study. 2019, 20 (3), 323–329. e2.

52. Ralph, S. J.; Espinet, A. J., Increased All-Cause Mortality by Antipsychotic Drugs: Updated Review and Meta-Analysis in Dementia and General Mental Health Care. Journal of Alzheimer’s disease reports 2018, 2 (1), 1–26.

53. Enache, D.; Fereshtehnejad, S.-M.; Kåreholt, I.; Cermakova, P.; Garcia-Ptacek, S.; Johnell, K.; Religa, D.; Jelic, V.; Winblad, B.; Ballard, C.; Aarsland, D.; Fastbom, J.; Eriksdotter, M., Antidepressants and mortality risk in a dementia cohort: data from SveDem, the Swedish Dementia Registry. 2016, 134 (5), 430–440.

54. Han, C.; Wang, S. M.; Lee, S. J.; Jun, T. Y.; Pae, C. U., Optimizing the Use of Aripiprazole Augmentation in the Treatment of Major Depressive Disorder: From Clinical Trials to Clinical Practice. Chonnam medical journal 2015, 51 (2), 66–80.

55. Hashioka, S.; McGeer, P. L.; Monji, A.; Kanba, S. J. C. N. S. A. i. M. C., Anti-inflammatory effects of antidepressants: possibilities for preventives against Alzheimer’s disease. 2009, 9 (1), 12–19.

56. Orgeta, V.; Tabet, N.; Nilforooshan, R.; Howard, R. J. J. o. A. s. D., Efficacy of antidepressants for depression in Alzheimer’s disease: systematic review and meta-analysis. 2017, 58 (3), 725–733.

57. Burke, S. L.; Maramaldi, P.; Cadet, T.; Kukull, W. J. I. j. o. g. p., Decreasing hazards of Alzheimer’s disease with the use of antidepressants: mitigating the risk of depression and apolipoprotein E. 2018, 33 (1), 200–211.

58. Correia, A. S.; Vale, N. J. P., Antidepressants in Alzheimer’s Disease: A Focus on the Role of Mirtazapine. 2021, 14 (9), 930.

59. Feinberg, S. M.; Fariba, K.; Saadabadi, A., Thioridazine. 2017.

60. Masopust, J.; Protopopová, D.; Vališ, M.; Pavelek, Z.; Klímová, B., Treatment of behavioral and psychological symptoms of dementias with psychopharmaceuticals: a review. Neuropsychiatric disease and treatment 2018, 14, 1211–1220.

61. Wei, H.; Li, A. P. J. D. M.; Disposition, Permeabilized Cryopreserved Human Hepatocytes as an Exogenous Metabolic System in a Novel Metabolism-Dependent Cytotoxicity Assay for the Evaluation of Metabolic Activation and Detoxification of Drugs Associated with Drug-Induced Liver Injuries: Results with Acetaminophen, Amiodarone, Cyclophosphamide, Ketoconazole, Nefazodone, and Troglitazone. 2022, 50 (2), 140–149.

62. Fabre, L. F.; Abuzzahab, F. S.; Amin, M.; Claghorn, J. L.; Mendels, J.; Petrie, W. M.; Dubé, S.; Small, J. G., Sertraline safety and efficacy in major depression: a double-blind fixed-dose comparison with placebo. Biological psychiatry 1995, 38 (9), 592–602.

63. Starkstein, S. E.; Mizrahi, R. J. E. r. o. n., Depression in Alzheimer’s disease. 2006, 6 (6), 887–895.

64. Lavergne, F.; Berlin, I.; Gamma, A.; Stassen, H.; Angst, J., Onset of improvement and response to mirtazapine in depression: a multicenter naturalistic study of 4771 patients. Neuropsychiatric disease and treatment 2005, 1 (1), 59–68.

65. Alam, A.; Voronovich, Z.; Carley, J. A., A review of therapeutic uses of mirtazapine in psychiatric and medical conditions. The primary care companion for CNS disorders 2013, 15 (5).

66. Stahl, S. M.; Pradko, J. F.; Haight, B. R.; Modell, J. G.; Rockett, C. B.; Learned-Coughlin, S., A Review of the Neuropharmacology of Bupropion, a Dual Norepinephrine and Dopamine Reuptake Inhibitor. Primary care companion to the Journal of clinical psychiatry 2004, 6 (4), 159–166.

67. Pandhare, A.; Pappu, A. S.; Wilms, H.; Blanton, M. P.; Jansen, M., The antidepressant bupropion is a negative allosteric modulator of serotonin type 3A receptors. Neuropharmacology 2017, 113 (Pt A), 89–99.

68. Farahani, A.; Correll, C. U. J. T. J. o. c. p., Are antipsychotics or antidepressants needed for psychotic depression? A systematic review and meta-analysis of trials comparing antidepressant or antipsychotic monotherapy with combination treatment. 2012, 73 (4), 1269.

69. Vázquez, G. H.; Bahji, A.; Undurraga, J.; Tondo, L.; Baldessarini, R. J., Efficacy and Tolerability of Combination Treatments for Major Depression: Antidepressants plus Second-Generation Antipsychotics vs. Esketamine vs. Lithium. 2021, 35 (8), 890–900.

