## Supplementary figures and images for "Combination of Antidepressants and Antipsychotics as A Novel Treatment Option for Psychosis in Alzheimer’s Disease"

### Supplemental Figure 1

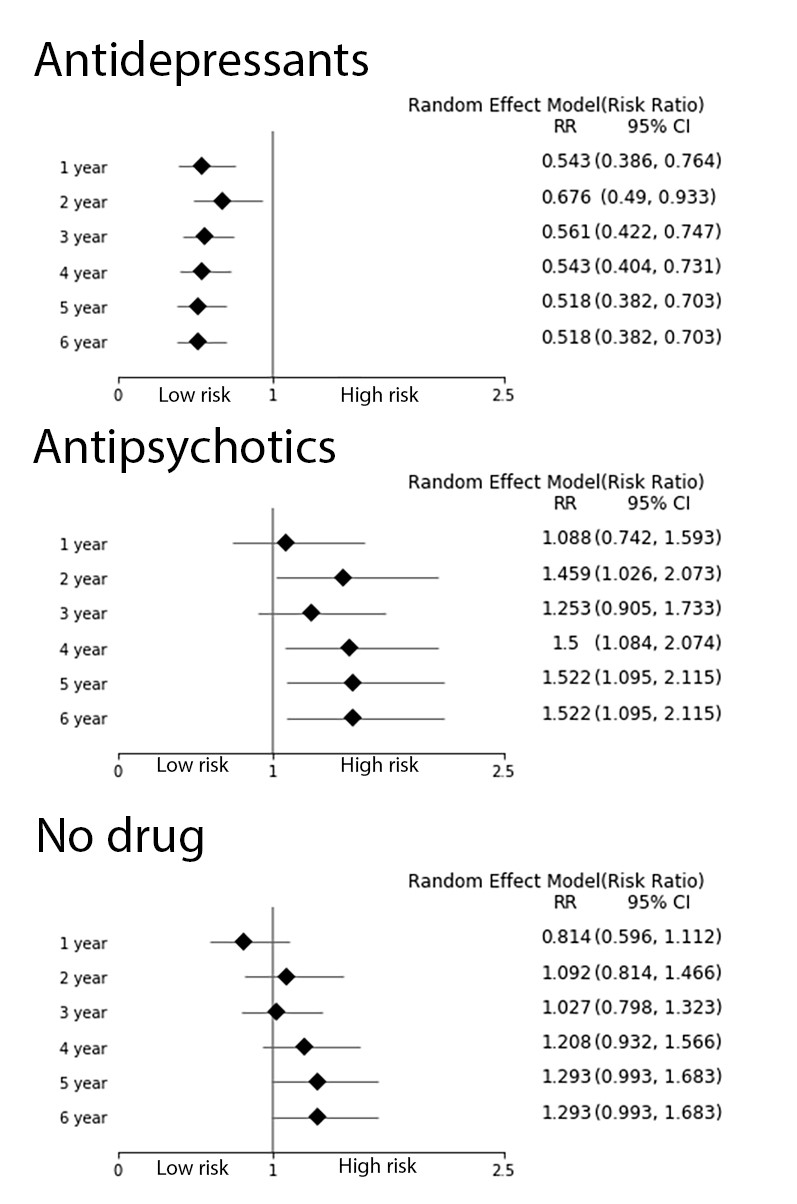

### Supplemental Figure 2

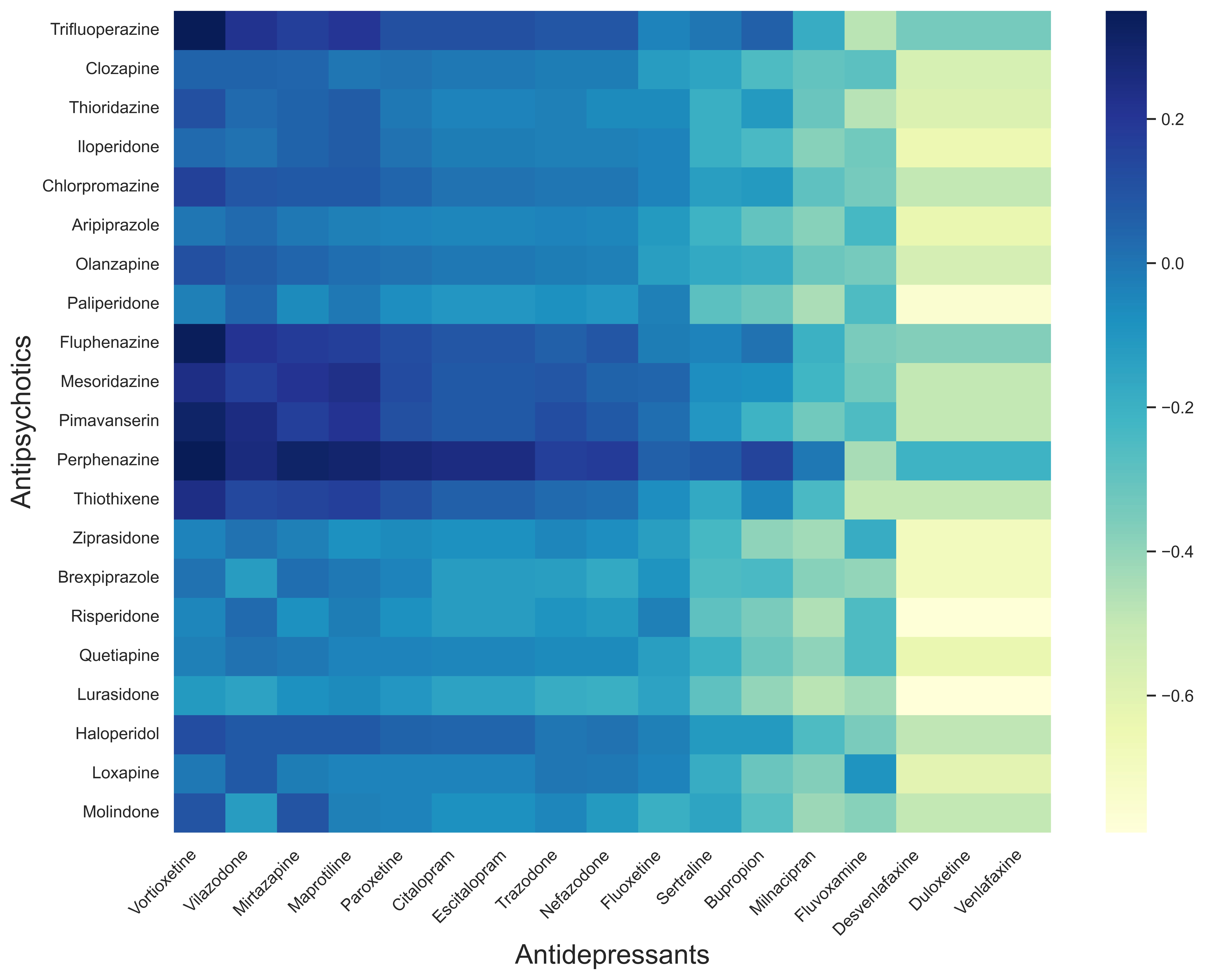
